# Gender differences in perceived health in relation to working conditions and socio-economic status in Spain, 2014-2017

**DOI:** 10.1101/2022.05.25.22275596

**Authors:** Amanda Godoy, Araceli Rojo, Luisa Delgado, José J Martín, M. Teresa Sánchez, M. Puerto López del Amo

## Abstract

A gender perspective was used to analyze how socio-economic status and per capita public health expenditure are associated to perceived health among the Spanish population between the years 2014 and 2017. Using multilevel methodologies (looking at year, individual, and region) and linear specification, the longitudinal microdata files from the Survey on Living Conditions were analyzed. The results point at low educational levels being a factor for worse perceived health among women, while for the same group income appears to have a protective influence. On the other hand, women are not negatively affected by unemployment, unlike men. Regional per capita public health expenditure is not associated with perceived health in either men nor women.

## Introduction

The theoretical framework concerning the social determinants of health posits that variables such as working status, educational attainment level, and socio-economic context are unequally linked to health [1-5]. In that frame of reference, gender appears as a particularly influential axis of inequality [6, 7].

Biological differences between sexes are insufficient to explain diverging health trends, which have been proven to be preventable and avoidable. Unlike sex, i. e. a set of biological attributes, the concept of gender derives from socially constructed cultural conventions, roles, and behaviors ascribed to people of the female, male, or other sexes [8]. Socially and culturally constructed gender norms determine the roles and opportunities afforded to all individuals and arise as strong structural determinants of health with major yet different implications for women and men [6, 9]. Such differences become problematic when they give rise to inequality and discrimination, which are particularly detrimental to women. Gender power dynamics, which are among the main causes of gender inequality, are also some of the strongest social determinants of health, with an undeniable effect on how people are born, grow up, live, work, get old, and finally die [10].

The main goal of the present work is to analyze, from a gender perspective, the association between perceived health and socio-economic status. Additionally, this study examines the extent to which regional public health expenditure affects the perceived health of men and women in a given region. For the purposes of this study, socio-economic status is defined along three dimensions: educational level, economic status, and working status. A set of instrumental variables have been selected for each dimension in order to provide a well-rounded description of the position each individual occupies.

For the most part, the relevant literature consists of partial analyses of the relation between health and certain social and economic variables, such as the role of unemployment or poverty on health [11-17]. In contrast, the present study considers the link between socio-economic status and health from a global point of view, by simultaneously contemplating the influence of educational level, economic status, and the individual’s working status as the three key dimensions of general socio-economic status.

Educational level has been proven to be a major social determinant of health. Most analyses confirm that high educational attainment is commonly linked to improved health and increased longevity with good health, when comparisons are drawn with individuals with low levels of education [18-20]. However, the specifics of how the benefits of education are unevenly distributed among women and men are not sufficiently explained, and the conclusions attained are far from unanimous. Given that women are generally at a disadvantage in the distribution of socio-economic resources such as power, authority, earnings, household income, and general wealth, their health and survival might be more dependent on their education than in the case of men. This study examines whether the beneficial influence of education on health is particularly strong for women, and whether education by itself may suffice to overcome gender differences in perceived health.

Previous research has concluded that lower educational levels are particularly detrimental to the perceived health of women when compared with similarly characterized men [19, 21-23]. A recent analysis carried out in Spain among an adult population established that the association between low educational level and poor self-perceived health was particularly strong among women [20]. However, an analysis of the active population in Spain failed to find gender disparities concerning the effect of education on health, although it showed that women’s health is lower among less educated individuals, mainly due to job insecurity and the specific characteristics of households [18].

Some research on psychological welfare suggests that education is more beneficial to women than men [24, 25], maybe because the lack of other resources makes women particularly dependent on their own education to achieve certain levels of welfare. Thus, poorly educated women may suffer from more and worse health problems because they have less resources to draw from [25-28]. The theory of resource substitution, according to which the absence of one or several socio-economic resources may be offset by drawing more intensely from the existing ones, predicts that the benefits of education on health and survival are larger for women, with physical deterioration being more intensely reduced for women than for men as their educational level increases. Thus, the gender gap in physical status essentially vanishes as individuals reach college levels of education. In contrast, the theory of resource multiplication implies precisely the opposite, that education should be more beneficial to men than women as they are able to derive greater rewards from it in the labor market, such as increased authority and income [25, 26].

The critical link between women’s education and health has been emphasized even more by evidence concerning the discrimination faced by women who attempt to access the labor market and the gender pay gap, which has increased as a result of the latest financial recession [29].

Individual economic status is a major social determinant of health, with all evidence pointing at income levels, poverty, and inequality being associated with worsened perceived health [30]. Sarti et al. (2019) found that in Italy decreased socio-economic status is associated to poorer self-perceived health [31], while Akanni et al. (2022) concluded that, in Germany, the increase and stability of household income had beneficial effects on health [32]. Income inequality has increased sharply over the last few decades, which may have contributed to perpetuate or exacerbate health disparities. In turn, poor health reduces income, which creates a negative feedback loop commonly known as the health-poverty trap [33]. The financial crisis modified working conditions and levels of employment, increasing job insecurity and decreasing wage income, which raised the share of those at risk of poverty and social exclusion [34]. In that regard, the World Health Organization deems poverty to be the strongest determinant of poor health at the individual level [35], with women being particularly vulnerable in that regard as the effects of want are added to gender inequalities and the so-called feminization of poverty, which affects women more commonly and intensely than men [36]. Generally speaking, poor living conditions and material deprivation have a detrimental effect on people’s health [37]. Such circumstances, particularly when prolonged in time, undermine their working status, erode their human and social capital, worsen their health, and brand them with stigma [38].

As far as social deprivation is concerned, the strong link between poverty and health is a widely established fact, notwithstanding the complex nature of both concepts and their relation [39]. As for material deprivation as a contributor to social deprivation, little empirical evidence has been gathered. Amendola et al. (2020) found, for the period 2007-2011, that although social deprivation in absolute terms had increased for men as well as women, the differences between both groups had been reduced: men had become more socially deprived, but women remained in the lead in the same regard [40]. The latest AROPE report (at risk of poverty and social exclusion) available for Spain [41] points at women being more intensely affected than men by material deprivation, as well as by the general AROPE score. These conclusions highlight the need to focus efforts on the female population.

Working status, and in particular the employed-unemployed dichotomy, has traditionally played a central role in the analysis of social inequalities concerning health. The intensity and magnitude of the Great Financial Crisis had a profound effect on the Spanish labor market, and the resulting landscape demands that we take a closer look at the experience of unemployment and how it is able to affect the health of women and men in a different way [42]. Previous analyses on the impact that unemployment has on health have revealed, in most cases, a negative influence [21, 43-45]. However, there is no consensus regarding the impact of unemployment on perceived health from a gender perspective. Norström et al. (2014) failed to find a clear, gender-determined inequality effect of unemployment on health [46]. Meanwhile, Drydakis (2015) found that, during the intense depression caused by the financial crisis in Greece, the detrimental effect of unemployment on health was stronger for women than for men [47]. In contrast with Drydakis, Antonakakis and Collins (2014) reached a different set of conclusions in their analysis of the Greek situation, as they found a starker effect of the financial recession and its austerity measures on male than on female suicide rates [48]. Other studies have similarly suggested that the effects of austerity, layoffs, and unemployment have had a stronger effect on the mental health of men than on that of women [49-52]. Along such lines, Calzón et al. (2017) found evidence of unemployment as a risk factor for bad self-rated health among men, but not women. However, the same study linked lower income levels to bad self-rated health to a greater extent in the case of women [22]. As for France, Ronchetti and Terriau (2019) did not find a negative link between long-term unemployment and self-perceived health for men nor for women [53]. Buffel et al. (2015), applying a multilevel framework to data extracted from the European Social Survey (2006 and 2012 rounds), looked into the relation between unemployment and mental health across 20 European countries, and found a larger increase in the likelihood of undergoing depression for men than for women [54]. In light of these studies, it becomes necessary to analyze how such effects may be modified by periods of economic recovery while keeping in mind the singularities of the Spanish economy, characterized by one of the highest unemployment rates in the European Union and a largely unstable and insecure labor market in which women experience higher rates of unemployment [34].

Lastly, we carried out an analysis of regional per capita health expenditure and perceived health, accounting for the significant differences in this variable across regions. The Public Health Expenditure Statistical Report published by the Spanish Ministry of Health (Estadística de Gasto Sanitario Público, 2022), reveals a 48% gap between the highest spending region (Basque Country) and the lowest (Andalusia) [55]. The differences are significant: even accounting for the Basque Country’s disparate legislative and administrative framework, the differences amount to 38%, only slightly less elevated. Among low-income countries, higher health expenditure seems to be linked to significant improvements in health status, which suggests that public policy may make a big difference in this regard [56, 57]. However, among high-income countries, additional health expenditure increases appear to be largely unrelated to health status improvements, which implies that increases in expenditure alone do not suffice to improve health to a significant degree [56-58]. Other studies, such as that carried out by Heijink et al. (2013) for 14 Western countries, did find a statistically significant link between health expenditure and avoidable mortality [59]. Nixon and Ulmann (2006) examined the relation between health care expenditure and health outcomes (average life expectancy at birth and infant mortality rate) to conclude that, although health expenditure has contributed significantly to ameliorate infant mortality, it has only caused marginal improvements to life expectancy in EU countries during the period under analysis [60]. Some researchers have been able to identify that women’s educational attainment levels, technological improvements, per capita income, unequal distributions of income, and certain cultural differences are behind some of the strongest improvements in health outcomes, far beyond increases in health expenditure [61, 62].

To summarize, in Spain as well as internationally, the available evidence concerning the link between socio-economic status, gender, and perceived health is far from conclusive or unanimous. The present study takes a wide longitudinal database to explore how health gender differences are affected by educational attainment level, economic status, and working status. The results of this analysis may be useful to suggest specific public policies with the potential to reduce health status gaps caused by the disparate social and economic status of women and men. Additionally, the following pages also looks into how the differences in public health expenditure of each region relate to the perceived health status of their citizens.

## Materials and methods

### Scope of the study

A database was built using the microdata files from the Survey on Living Conditions (2014-2017), comprising 17,027 individuals aged between 16 and 60, and 41,111 observations [63]. Per capita public health expenditure was extracted from the Public Health Expenditure Statistical Report published by the Spanish Ministry of Health *(Estadística de Gasto Sanitario Público)* [55].

### Data analysis

Taking into account the chronological and hierarchical structure of the data, a multilevel (three-level) longitudinal model was estimated. The first level was the year, the second the individual, and the third level was the region. Analytically:

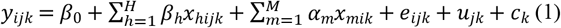

where *y*_*ijk*_ is the dependent variable (perceived health), taking values 1-5, for the year *i*, being i= 2014, …, 2017; the individual *j*, being *j* = 1,…,17,027; and region *k*, being *k* = 1,…,17. *β*_0_ represents the independent term; *x*_*hijk*_ are the individual explanatory variables; *j* and *β*_*h*_ their associated coefficients, with *h* = 1,…,*H*; *x*_*mik*_ are the explanatory variables at the ecological level, *k* and *α*_*m*_ their associated coefficients, with *m* = 1,…,*M*. The error term divides the unexplained part in three, one for each hierarchical level: *c*_*k*_ at the ecological level, *u*_*jk*_ for the individual, and *e*_*ijk*_ for each year.

Multilevel models have been proven to be a good fit for hierarchical structures that include several levels of information, and in which individuals share certain features by virtue of belonging to the same higher level (the region). They are also a good choice when repeated measures exist, given that they allow for variance to be estimated at each level. Multilevel models thus avoid both the ecological fallacy (in which aggregated data are interpreted at the individual level) and the atomistic fallacy (in which individual data are interpreted at the aggregated level) [64]. The software employed to analyze the data was Stata 15.

The dependent variable, perceived health, takes values 1-5, with 1 for very good health and 5 for very bad health. Perceived health is a proxy variable for morbi-mortality, commonly employed in health and living conditions surveys. It is one of the indicators of choice used to monitor gender inequalities and their determinants in matters of health [65-67].

Individually, the variables of interest are those characterizing the socio-economic status of subjects: educational attainment level, economic status, and working status. Educational level is defined as the amount of formal instruction successfully completed, and is divided into the primary, secondary, and college level.

Economic status has been characterized by means of four instrumental variables. The first is income, on the one hand, and on the other are three indicators that describe social deprivation: the AROPE score and two of its components, severe material deprivation and low work intensity in the household (LWIH).

Income was measured using the equivalent available per capita income of households at constant levels, for the last year of the per capita income series and logarithmically transformed.

The risk of poverty and social exclusion was measured through the AROPE score. This indicator expands on the traditional relative poverty score by combining it with severe material deprivation and low work intensity in the household. This provides a multidimensional picture of poverty and social exclusion. The AROPE score thus comprises three sub-indicators representing three distinct population sets: individuals at risk of poverty, individuals suffering from severe material deprivation, and individuals living in households characterized by low work intensity. Whenever one such indicator is present, an individual can be safely described as being at risk of poverty or social exclusion [41]. The variable *low work intensity* refers to individuals living in households in which those of working age did work less than 20% of their full potential in the year before the survey. As for severe material deprivation, this describes the share of the population living in households lacking in at least four out of nine specific quality-of-life items [68].

Finally, working status has been characterized using six categories: employed, unemployed, student, homemaker, retired, and other inactive status.

Age and chronic illness (dichotomous variable) were used as control variables.

For the third level, which deals with the region, per capita public health expenditure was used. A one-year delay was introduced in order to better reflect any potential link with the perceived health of residents in the region.

All monetary variables, such as income and health expenditure, have been expressed using 2017 prices.

## Results

Table 1 shows the frequency of each variable in the model, split by gender.

**Table 1.** Descriptive statistical values, according to gender, of the variables used to measure the relation of socio-economic and working status with perceived health for Spain between 2014 and 2017.

|  |  | WOMEN |  |  |  |  | MEN |  |  |  |  |
| --- | --- | --- | --- | --- | --- | --- | --- | --- | --- | --- | --- |
|  |  | 2014 | 2015 | 2016 | 2017 | Total | 2014 | 2015 | 2016 | 2017 | Total |
| Categorical variables |  | n=2668<br>% | n=4997<br>% | n=7104<br>% | n=6087<br>% | n=20856<br>% | n=2573<br>% | n=4817<br>% | n=6966<br>% | n=5899<br>% | n=20255<br>% |
| Perceived health | 1 | 19.72 | 18.69 | 17.23 | 22.41 | 19.41 | 21.14 | 20.78 | 20.13 | 24.17 | 21.59 |
|  | 2 | 60.31 | 63.18 | 64.20 | 61.00 | 62.52 | 63.04 | 62.40 | 63.58 | 60.91 | 62.45 |
|  | 3 | 15.74 | 14.45 | 15.05 | 13.34 | 14.49 | 12.16 | 13.29 | 13.36 | 12.04 | 12.81 |
|  | 4 | 3.26 | 2.94 | 2.83 | 2.73 | 2.88 | 2.99 | 2.70 | 2.41 | 2.29 | 2.52 |
|  | 5 | 0.97 | 0.74 | 0.69 | 0.53 | 0.69 | 0.66 | 0.83 | 0.52 | 0.59 | 0.63 |
| Chronic illness | 1 | 23.61 | 24.13 | 23.10 | 21.24 | 22.87 | 21.53 | 23.87 | 21.60 | 19.33 | 21.47 |
|  | 0 | 76.39 | 75.87 | 76.90 | 78.76 | 77.13 | 78.47 | 76.13 | 78.40 | 80.67 | 78.53 |
| Educational level | Primary | 12.29 | 10.51 | 9.87 | 8.00 | 9.79 | 13.84 | 11.86 | 11.25 | 9.26 | 11.14 |
|  | Secondary | 54.39 | 55.03 | 54.89 | 54.51 | 54.75 | 58.09 | 58.39 | 58.73 | 58.92 | 58.63 |
|  | College | 33.32 | 34.46 | 35.24 | 37.49 | 35.46 | 28.07 | 29.75 | 30.02 | 31.82 | 30.23 |
| Working status | Employed | 51.37 | 53.07 | 54.30 | 56.70 | 54.33 | 60.70 | 64.50 | 64.45 | 67.46 | 64.87 |
|  | Unemployed | 21.62 | 20.50 | 19.27 | 15.75 | 18.83 | 23.22 | 19.44 | 18.03 | 15.45 | 18.27 |
|  | Student | 10.46 | 11.04 | 11.20 | 11.06 | 11.01 | 10.42 | 10.31 | 11.67 | 10.91 | 10.96 |
|  | Homemaker | 12.76 | 12.01 | 11.56 | 12.12 | 11.97 | 0.07 | 0.08 | 0.14 | 0.15 | 0.12 |
|  | Retired | 0.25 | 0.15 | 0.26 | 0.51 | 0.31 | 0.87 | 0.76 | 0.87 | 1.30 | 0.97 |
|  | Other inactive | 3.54 | 3.23 | 3.50 | 3.87 | 3.55 | 4.72 | 4.91 | 4.84 | 4.73 | 4.81 |
| Severe material deprivation | 1 | 6.35 | 4.67 | 4.30 | 3.05 | 4.29 | 5.70 | 4.59 | 3.98 | 3.10 | 4.09 |
|  | 0 | 93.65 | 95.33 | 95.70 | 96.95 | 95.71 | 94.30 | 95.41 | 96.02 | 96.90 | 95.91 |
| Low work intensity in the household (LWIH) | 1 | 20.90 | 17.11 | 16.16 | 14.39 | 16.48 | 19.44 | 16.65 | 15.20 | 12.94 | 15.43 |
|  | 0 | 79.10 | 82.89 | 83.84 | 85.61 | 83.52 | 80.56 | 83.35 | 84.80 | 87.06 | 84.57 |
| AROPE | 1 | 35.24 | 31.87 | 30.91 | 29.39 | 31.25 | 33.60 | 30.46 | 29.10 | 26.92 | 29.36 |
|  | 0 | 64.76 | 68.13 | 69.09 | 70.61 | 68.75 | 66.40 | 69.54 | 70.90 | 73.08 | 70.64 |
| Continuous variables |  | mean (SD) |  |  |  |  | mean (SD) |  |  |  |  |
| Age | Age | 40.53<br>(12.43) | 40.61<br>(12.58) | 41.07<br>(12.68) | 41.33<br>(12.83) | 40.96<br>(12.67) | 39.72<br>(12.50) | 40.21<br>(12.65) | 40.41<br>(12.79) | 40.78<br>(12.93) | 40.38<br>(12.77) |
| Household income | nldeflat_equivincome | 9.41<br>(0.74) | 9.45<br>(0.76) | 9.49<br>(0.76) | 9.50<br>(0.75) | 9.47<br>(0.76) | 9.44<br>(0.71) | 9.47<br>(0.74) | 9.51<br>(0.72) | 9.53<br>(0.72) | 9.50<br>(0.72) |
| Per capita health expenditure | healthexp/pc | 1293.42<br>(150.23) | 1385.97<br>(146.46) | 1412.39<br>(159.10) | 1410.20<br>(156.12) | 1390.16<br>(158.89) | 1288.63<br>(149.44) | 1387.39<br>(145.69) | 1412.28<br>(159.10) | 1411.84<br>(156.43) | 1390.47<br>(159.15) |

Source: Prepared by the authors using data from Instituto Nacional de Estadística (INE) (2020). Carencia Material. Carencia Material Severa [Material Deprivation. Severe Material Deprivation]. Recovered from https://www.ine.es/ss/Satellite?L=es_ES&c=INESeccion_C&cid=1259925456180&p=1254735110672&pagename=ProductosYServicios%2FPYSLayout&param1=PYSDetalle&param3=1259924822888, Instituto Nacional de Estadística (INE) (2022). Encuesta de condiciones de vida. Resultados. Available from: https://www.ine.es/dyngs/INEbase/es/operacion.htm?c=Estadistica_C&cid=1254736176807&menu=resultados&idp=1254735976608#!tabs-1254736195153 and Ministerio de Sanidad del Gobierno de España (2022) Estadística de Gasto Sanitario Público (EGSP) 2019: Principales resultados. Available from: https://www.msbs.gob.es/estadEstudios/estadisticas/docs/EGSP2008/egspPrincipalesResultados.pdf.

According to the analysis, the share of individuals describing their health as fair, bad, or very bad has increased between 2014 and 2016, and only slightly decreased in 2017. Such temporary respite affected women and men alike. As for the variables of interest, the prevailing educational level for women and men in the whole series under analysis is secondary education, followed by college-level studies. Regarding economic status, both women and men enjoyed higher income levels in the last year under analysis: 2017. The three indicators used to describe social deprivation successively decreased over the 2014-2017 period for both genders alike. Finally, and as far as working status is concerned, for the period under analysis a sustained increased was observed in the number of employed individuals, with a corresponding decrease in unemployment rates, also for women as well as men. The working status category displaying the largest gap is that of homemakers, which includes almost 12% of women but only 0.12% of men.

Table 2 displays the results of estimations.

**Table 2.** Results of applying the random intercept linear multilevel model to the exploration of associations between individual working and socio-economic status and perceived health.

|  | WOMEN |  |  | MEN |  |  |
| --- | --- | --- | --- | --- | --- | --- |
|  | Null model | Explanatory model |  | Null model | Explanatory model |  |
| Variable | Coefficient (SE) | p value | 95% Conf. Interval | Coefficient (SE) | p value | 95% Conf. Interval |
| **Constant** | 1.527 (0.123) | 0.000 | (1.287 1.768) | 1.189 (0.220) | 0.000 | (0.757 1.620) |
| **Age** | 0.012 (0.001) | 0.000 | (0.010 0.013) | 0.013 (0.001) | 0.000 | (0.011 0.014) |
| **Chronic illness** | 0.766 (0.014) | 0.000 | (0.738 0.794) | 0.701 (0.022) | 0.000 | (0.659 0.744) |
| **Working status** |  |  |  |  |  |  |
| - **Employed** | Reference |  |  | Reference |  |  |
| - **Unemployed** | 0.013 (0.026) | 0.617 | (-0.038 0.065) | 0.069 (0.016) | 0.000 | (0.037 0.102) |
| - **Student** | -0.062 (0.019) | 0.001 | (-0.099 -0.024) | -0.034 (0.014) | 0.015 | (-0.061 -0.006) |
| - **Homemaker** | 0.001 (0.012) | 0.934 | (-0.023 0.026) | 0.045 (0.157) | 0.776 | (-0.264 0.353) |
| - **Retired** | 0.115 (0.097) | 0.234 | (-0.074 0.305) | 0.164 (0.062) | 0.008 | (0.043 0.285) |
| - **Other inactive** | 0.428 (0.045) | 0.000 | (0.339 0.516) | 0.563 (0.037) | 0.000 | (0.490 0.635) |
| **Education level** |  |  |  |  |  |  |
| - **Primary** | 0.070 (0.013) | 0.000 | (0.044 0.097) | 0.028 (0.017) | 0.090 | (-0.004 0.061) |
| - **Secondary** | Reference |  |  | Reference |  |  |
| - **College** | -0.077 (0.012) | 0.000 | (-0.100 -0.053) | -0.098 (0.019) | 0.000 | (-0.135 -0.060) |
| **Social deprivation:** |  |  |  |  |  |  |
| - **LWIH** | 0.053 (0.025) | 0.034 | (0.004 0.103) | 0.036 (0.014) | 0.014 | (0.007 0.064) |
| - **Severe material deprivation** | 0.129 (0.025) | 0.000 | (0.081 0.178) | 0.158 (0.029) | 0.000 | (0.101 0.215) |
| - **AROPE** | 0.046 (0.024) | 0.052 | (-0.000 0.093) | 0.058 (0.016) | 0.000 | (0.027 0.089) |
| - **nl_def_equiv. income** | -0.028 (0.010) | 0.006 | (-0.048 -0.008) | -0.007 (0.015) | 0.617 | (-0.037 0.022) |
| **Per capita health expend.** | 0.000 (0.000) | 0.404 | (-0.000 0.000) | 0.000 (0.000) | 0.208 | (-0.000 0.000) |

LWIH (Low Work Intensity in the Household) AROPE (At Risk of Poverty and Social Exclusion)
Source: Prepared by the authors using data from Instituto Nacional de Estadística (INE) (2020). Carencia Material. Carencia Material Severa [Material Deprivation. Severe Material Deprivation]. Recovered from https://www.ine.es/ss/Satellite?L=es_ES&c=INESeccion_C&cid=1259925456180&p=1254735110672&pagename=ProductosYServicios%2FPYSLayout&param1=PYSDetalle&param3=1259924822888, Instituto Nacional de Estadística (INE) (2022). Encuesta de condiciones de vida. Resultados. Recovered from https://www.ine.es/dyngs/INEbase/es/operacion.htm?c=Estadistica_C&cid=1254736176807&menu=resultados&idp=1254735976608#!tabs-1254736195153 and Ministerio de Sanidad del Gobierno de España (2022) Estadística de Gasto Sanitario Público (EGSP) 2019: Principales resultados. Recovered from: https://www.mscbs.gob.es/estadEstudios/estadisticas/docs/EGSP2008/egspPrincipalesResultados.pdf.

Regarding the variables of interest that characterize socio-economic status, for women statistically significant associations appeared with educational attainment level (for education-related variables); income, severe material deprivation, and living in a household with low work intensity (for economic status-related variables); and belonging to the *other inactive* or *student* categories (for working status-related variables). Having successfully completed only a primary-level education is a risk factor for women (β= 0.070, p=0.00), whereas having received a college-level education has a protective effect (β= - 0.077, p=0.00).

The following results have been observed with respect to the economic status of women: Income is a protective factor (β= - 0.028, p < 0.05), whereas severe material deprivation (β= 0.129, p = 0.00), living in a low work intensity household (β= 0.053, p < 0.05) and the AROPE rating (β= 0.046, p < 0.10) are definite risk factors.

Finally, as regards working status, only belonging to the *other inactive* (β= 0.428, p=0.00) o *student* categories (β= - 0.062, p < 0.05) yielded statistically significant results. It is worth pointing out that all other working status categories lacked statistical significance, notably being unemployed (β= 0.013, p=0.617) and being a homemaker (β= 0.001, p=0.934).

The per capita public health expenditure variable was not statistically significant (β= 0.000, p=0.404) and therefore is not associated with better or worse perceived health among women.

In the case of men, the multilevel analysis shows that, as far as education is concerned, only having received a college-level education has a statistically significant protective effect (β= - 0.098, p=0.00). As for economic status, risk factors include severe material deprivation, living in a low work intensity household, and the AROPE rating. Finally, as regards working status, unlike in the case of women unemployment and retirement appeared as risk factors.

The analysis of the economic status of men yielded the following results:

Income did not reach statistical significance (β= - 0.007, p = 0.617), whereas severe material deprivation (β= 0.158, p = 0.000), living in a household with low work intensity (β= 0.036, p < 0.05), and the AROPE score (β= 0.058, p = 0.000) appeared as risk factors. Finally, and concerning working status, being unemployed (β= 0.069, p = 0.000) and retired (β= 0.164, p < 0.05) were definite risk factors for men.

The public health expenditure variable was not statistically significant (β= 0.000, p=0.208), which implies that regional per capita public health expenditure is not associated with the improved or worsened perceived health of men.

To summarize, the main differences observed by gender are the following:

– In matters of educational level, for women having received only a primary education worsened their perceived health by 0.07 points (in contrast with being in possession of a secondary education). Conversely, for men a primary education level was not a risk factor for health.
– As regards economic status, income was a protective factor for the good health of Spanish women. A 10% increase in income improves the health of women by two percentage points. In contrast, for men income did not have any noticeable effect.
– As for working status, the perceived health of Spanish women does not appear to be affected by unemployment, whereas for men it decreases their self-rated health status. Unemployed men were, on average, 0.07 percentage points below their employed peers in perceived health. Similarly, being retired does not have any effect in the case of women, but for men it worsens their self-perceived health. Retired men report a health status 0.16 percentage points below those who are employed.

Therefore, it may be stated that variables describing economic status, be they severe material deprivation, LWIH, or the AROPE score, have a deleterious effect on the health of both women and men.

In the analysis of gender differences, for women a low educational level is a risk factor and higher income has a protective effect, whereas retirement and unemployment are negatively associated with the health of men and men alone. Regional per capita public health expenditure is not statistically associated to the perceived health of women nor men.

## Discussion

The goal of this analysis was to provide a gender perspective on the association between perceived the health and socio-economic status of the Spanish population between the years of 2014 and 2017.

This multilevel analysis employed data from the Living Conditions Survey and from several regional sources, and its results suggest that, although there are certain common factors which affect the health of women and men alike, others have a differential effect. This points at strong gender differences as far as health and its socio-economic determinants are concerned.

Firstly, our results show that a low educational attainment level is a risk factor for women, but not for men. In agreement with previous studies [20-22, 57], we found that women’s health is further improved by the intrinsic rewards of education, as they have fewer other resources from which they can draw in the absence of a degree. Whenever women are able to complete an education, their health is improved, often at a higher rate than for men. This illustrates the point that women are especially reliant on their own education in order to thrive, and therefore improvements in education may reduce their health-damaging behaviors further than those of men [25]. This result suggests that public investments in women’s education translate to a reduction in health-related gender differences.

As for the effects of economic status, results show that income is a protective factor for the health of women, but not for men. Generally speaking, people with higher income levels enjoy improved health as they have access to the resources required to prevent and treat a variety of conditions and are therefore better equipped to face critical and stressful events throughout their lives [30]. Women, however, are still at an economic disadvantage, and for that reason higher income levels may be a stronger predictor of their good health than of men’s [69-71].

Regarding the three dimensions of social deprivation under analysis (severe material deprivation, low work intensity in the household, and the AROPE score), they appear to negatively affect the health of women and men alike, consistently with previous studies such as López del Amo et al. (2018) [19].

With respect to working status, unemployment does not have an effect on the perceived health of women, but it worsens that of men, as previously reported by Calzón et al. (2017) [22]. The myth of the nuclear family and the concept of manhood as a function of a man’s ability to provide for their family still lie behind currently prevailing notions of masculinity and patriarchy [50, 51, 72]. These misconceptions have failed to adapt to new landscapes of family and work life, which may lead to increased physical and mental health problems for men in times of economic turmoil and uncertainty [73]. As reported by the World Bank (2011) men see access to the labor market as their only avenue for empowerment, and any setback in that regard makes them vulnerable [49]. In that sense, it must be noted that the collapse of the building industry at the beginning of the financial crisis left many people unemployed for a long time, with male unemployment rates increasing at a much faster pace than female rates. An analysis of national health surveys shows that the mental health of such individuals was severely affected during that period. Lastly, observations of per capita public health expenditure failed to reveal any effect on the perceived health of women nor men. While it is widely understood that it should have a strong effect on the health status of individuals, in developed countries additional expenditure is often unrelated to improvements in perceived health, as previously described in the literature [56-58]. This does not discount the fact that public policy makers should always be on the lookout for new opportunities to improve health outcomes through regionally distributed health expenditure and efficient policies.

Furthermore, statistics on health outcomes may be improved by a reassignment of resources, from healthcare to social programs. This is particularly the case of policies which have been proven to have a strong effect on the social determinants analyzed, such as those of an educational nature. Education is able to create human capital and promote the pre-distribution of income and wealth, remaining to this day the variable with the strongest explanatory power regarding health status and gender differences.

To summarize, active employment policies, programs aimed at reducing poverty, and initiatives that complement low income and improve educational opportunities may play a central role in alleviating poverty and improving the population’s health.

These results are particularly relevant in facing the current post-COVID crisis, as they may help guide public policy in matters of education and income re-distribution with the goal of promoting the post-pandemic recovery of our societies.

## Data Availability

Longitudinal Life Conditions Survey 2014-2017. Gender differences in perceived health in relation to working conditions and socio-economic status in Spain http://hdl.handle.net/10481/74838

## Acknowledgments

This work was supported by the Health Department of the Regional Andalusian Government (2017–2019) under Grant PI-0457-2016.

We thank Professor Manuel Correa for his suggestions and contributions.

## Declaration of Interest

None

## Notes

### Competing Interest Statement

The authors have declared no competing interest.

### Funding Statement

This work was supported by the Health Department of the Regional Andalusian Govern-ment (2017–2019) under Grant PI-0457-2016.

